# Transmission dynamics and control measures of COVID-19 outbreak in China: a modelling study

**DOI:** 10.1101/2020.07.09.20150086

**Authors:** Xu-Sheng Zhang, Emilia Vynnycky, Andre Charlett, Daniela de Angelis, Zhengji Chen, PHE COVID-19 modelling group, Wei Liu

**Affiliations:** Centre for Infectious Disease Surveillance and Control, National Infection Service, Public Health England, 61 Colindale Avenue, London NW9 5EQ, UK; Medical Research Council Centre for Outbreak Analysis and Modelling, Department of Infectious Disease Epidemiology, Imperial College Faculty of Medicine, Norfolk Place, London W2 1PG, UK; TB Modelling Group, TB Centre, Centre for Mathematical Modelling of Infectious Diseases and Faculty of Epidemiology and Population Health, London School of Hygiene and Tropical Medicine, London, UK; Medical Research Council Biostatistics Unit, University Forvie Site, Robinson Way, Cambridge CB2 0SR, UK; School of Public Health, Kunming Medical University, Kunming, Yunnan, P. R. China

**Keywords:** COVID-19, mainland China, modelling, mortality, transmissibility

## Abstract

COVID-19 is reported to have been effectively brought under control in China at its initial start place. To understand the COVID-19 outbreak in China and provide potential lessons for other parts of the world, in this study we combine a mathematical modelling with multiple datasets to estimate its transmissibility and severity and how it was affected by the unprecedented control measures. Our analyses show that before 29^th^ January 2020, the ascertainment rate is 6.9%(95%CI: 3.5 – 14.6%); then it increased to 41.5%(95%CI: 30.6 – 65.1%). The basic reproduction number (*R*_0_) was 2.23(95%CI: 1.86 – 3.22) before 8^th^ February 2020; then it dropped to 0.04(95%CI: 0.01 – 0.10). This estimation also indicates that the effect on transmissibility of control measures taken since 23^rd^ January 2020 emerged about two weeks late. The confirmed case fatality rate is estimated at 4.41%(95%CI: 3.65 – 5.30%). This shows that SARS-CoV-2 virus is highly transmissible but less severe than SARS-CoV-1 and MERS-CoV. We found that at the early stage, the majority of *R*_0_ comes from the undetected infected people. This implies that the successful control in China was achieved through decreasing the contact rates among people in general populations and increasing the rate of detection and quarantine of the infected cases.

## Introduction

An outbreak of severe pneumonia, an infectious disease now named as COVID-19, was reported by Chinese authorities in December 2019. The aetiological agent, SARS-CoV-2, a novel coronavirus, was isolated by Chinese authorities on 7 January 2020 and reported by WHO on 9^th^ January 2020. The first case of COVID-19 was reported to have symptom onset on 1^st^ December 2019 in Wuhan city of Huber province, China^1^, and the virus was quickly spread to other parts of China^2,3^ and the first case was reported outside of China on 13^th^ January 2020 (see Table 1). Because of the rapid spread of the disease, WHO announced the outbreak of COVID-19 as a “public health emergency of international concern” on 30^th^ January 2020 and assessed COVIOD-19 as a pandemic on 11^th^ March 2020^4^. Up to 25^th^ March 2020, 405,742 cases and 18,791 fatalities associated with COVID-19 have been reported globally. Of these, 81,218 cases and 3,281 fatalities have been reported in mainland China. Up to 10^th^ April 20020 (25 days late), the global numbers increase hugely to 1,521,252 cases and 92,798 deaths while the corresponding numbers in mainland China are 81,953 cases and 3,339 deaths. These numbers of COVID-19 cases and fatalities indicate that the outbreaks outside of China develop quickly, whereas the daily reported number of new cases in mainland China remained very low. This situation of mainland China can be attributed to the draconian and rapid control measures taken in China since late January 2020. The control measures started from the lockdown of the epicentre Wuhan city on 23^rd^ January 2020 and then extended to the whole of mainland China^3^. Extreme response measures included the complete shutdown and isolation of whole cities, cancellation of Chinese New Year celebrations, and prohibition of attendance at school and work, broadcast of critical information (e.g., promoting hand washing, mask wearing, and care seeking) with high frequency through multiple channels, massive mobilization of health and public health personnel as well as military medical units, rapid construction of entire hospitals for patients with severe symptoms, and reconstruction of shelters for patients with none or mild symptoms. Control measures integrate travel bans and restrictions, contact reductions and social distancing, screening and contact tracing, early case identification and isolation^2,5^ to curb the epidemic. The massive vigorous actions taken by the Chinese government and Chinese people have slowed down the epidemic of COVID-19 and now the spread of the disease has been brought under control in China.

**Table 1.** Timeline of outbreak in mainland China.

| date | Event |
| --- | --- |
| November 2019 | Several pneumoniae of unknown aetiology were discovered in Wuhan city, Hubei province, China |
| 01/12/2019 | Symptom onset of the first case reported |
| 31/12/2019 | An alert was issued by the Wuhan Municipal Health Commission and a rapid response team was sent to Wuhan by Chinese Centre for Disease Control and Prevention (China CDC)<br>The WHO China Country Office was informed of 27 cases of pneumonia of unknown aetiology detected in Wuhan city, Hubei Province of China. |
| 01/01/2020 | Wuhan's Huanan Seafood Wholesale Market, the most probable index source of zoonotic COVID-19 infection, was shut down and disinfected<br>China emergency response team was constructed. |
| 02/01/2020 | The first 41 confirmed cases were identified as laboratory-confirmed COVID-19 in Wuhan city |
| 03/01/2020 | A total of 44 patients with pneumonia of unknown aetiology have been reported to World Health Organisation (WHO) by Chinese authorities.<br>Chinese National Health Commission (CNHC) issued Diagnosis and Treatment plan for pneumonia of unknown aetiology |
| 04/01/2020 | One exported case from Wuhan city into Weng Zhou, Zhejiang Province, China |
| 05/01/2020 | A total of 59 patients with pneumonia of unknown aetiology have been reported to WHO by Chinese authorities |
| 07/01/2020 | The causative pathogen was identified as a novel coronavirus and genomic characterisation and test method development ensued. |
| 12/01/2020 | Notification of the novel coronavirus and its sequence was made to WHO |
| 13/01/2020 | First exported case was reported in Thailand |
| 17/01/2020 | A total of 62 patients with pneumonia of unknown aetiology have been reported |
| 19/01/2020 | A total of 198 patients with pneumonia of unknown aetiology have been reported |
| 20/01/2020 | Person-to-person transmission was confirmed and announced in China.<br>China's "National Infectious Disease Law" was amended to make COVID-19 a class B notifiable disease and its "Frontier Health and Quarantine Law" was amended to support the COVID-19 outbreak response effort. By doing this, Chinese law required all cases to be immediately reported to China's Infectious Disease Information System.<br>CNHC started daily situation report of COVID-19 on their official website. |
| 23/01/2020 | Lockdown of the epicentre Wuhan city, limiting mobility of people in and out of Wuhan, and then the control measures expanded quickly to the neighbouring areas |
| 25/01/2020 | The Chinese government raised the response level of the “Preparedness and Response Plan for Novel Infection Disease of Public Health Significance” to the Emergency level, based on the assessment that the risk of health impact caused by COVID-19 on the local population is high and imminent.<br>Chinese government announced its highest-level commitment and mobilized all sectors to respond to the epidemic and prevent further spread of COVID-19 |
| 31/01/2020 | WHO announced the outbreak of novel coronavirus as a “public health emergency of international concern” |
| 12/02/2020 | Definition of confirmed cases for Hubei province was changed to include clinically diagnosed cases or PRC tested positive. The extra high daily number of cases 15152 was reported from Hubei province.<br>On 29/02/2020 this changed back to “clinically diagnosed plus PCR tested positive “ |
| 11/03/2020 | WHO assessed COVID-19 as a pandemic |
| 17/04/2020 | CNHC raised the total death number in Wuhan city from 2579 to 3869 |

As a novel coronavirus, it is important to understand its transmissibility and severity from the data of the outbreak and to judge how effectively the control measures taken in China have helped stop the spread of SARS-CoV-2 virus in mainland China. This understanding should provide some critical information for controlling COVID-19 outbreaks in other parts of the world and for its potential re-emergence in future. In particular, exploring the main driver of the spread will provide practically useful information for policy makers to make effective non-pharmaceutical interventions at its early occurrence. To estimate the early dynamics of transmission in Wuhan, Kucharski et al^6^ fitted a stochastic transmission dynamic model to four datasets up to 11^th^ February 2020: daily number of new internationally exported cases; daily number of new cases in Wuhan with no market exposure; daily number of new cases in China; and proportion of infected passengers on evacuation flights. They estimated that the median daily reproduction number (*R*_t_) in Wuhan declined from 2.35 (95% CI: 1.15–4.77) 1 week before travel restrictions were introduced on 23^rd^ January 2020, to 1.05 (95% CI: 0.41– 2.39) 1 week after. Many other modelling studies ^2,3,7,8^ also used the confirmed case data at the early stage to model and estimate the spread dynamics and control of COVID-19 outbreak in China. As the number of new confirmed case in mainland China reached a peak only after 17^th^ February 2020 and other relevant data such as fatalities and hospital discharges are becoming available, a further study including more data streams is needed to show the overall effectiveness of the integrated control measures executed in the whole nation. Based on the observed data associated with the confirmed cases, death and recovery that were published by Chinese National Health Commission, in this study we propose a Synthesis model (Figure 1) which covers both transmission dynamics of SARS-CoV-2 virus and disease reporting process of COVID-19 to estimate the transmissibility and severity of COVID-19 in China and the impact on the spread of COVID-19 of control measures the Chinese government has taken from late January 2020.

**Figure 1.**
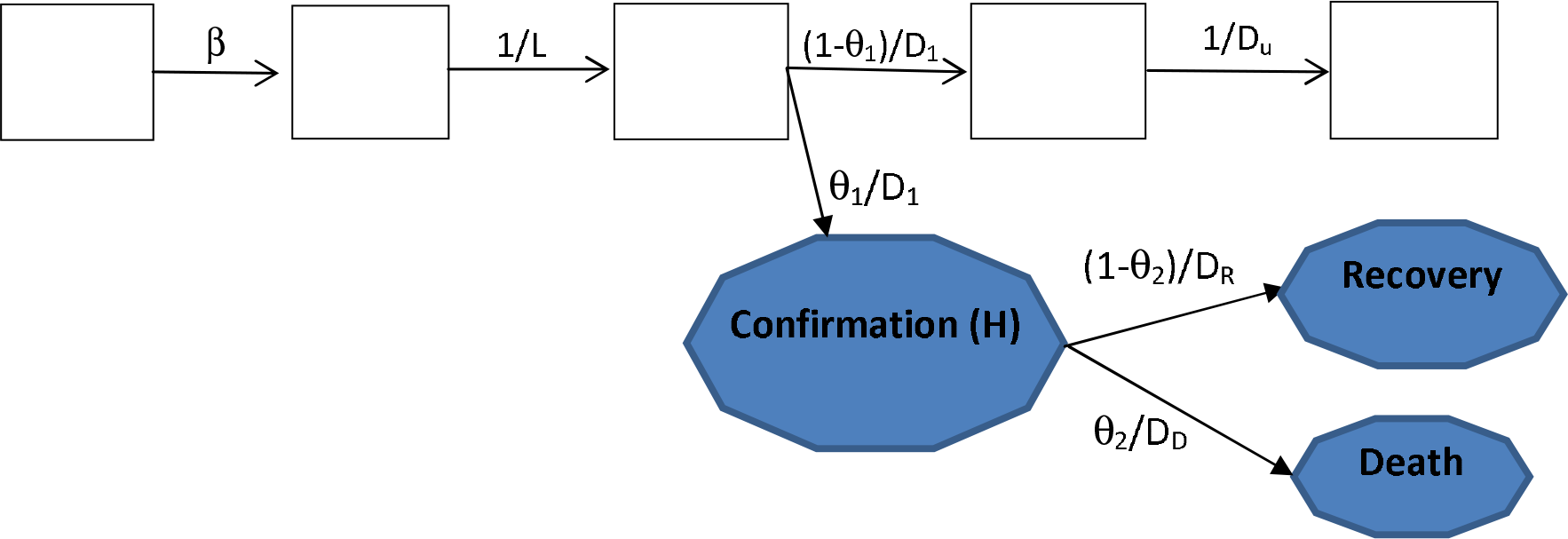
Flow chart of synthesis model: transmission dynamics and disease reporting processes. The five rectangle boxes represent the hidden transmission dynamics process and the three shaded polygons represent the quantities upon which observations were made.

## Results

The estimates of the model parameters are shown in Table 2. The detection and reporting rate (or ascertainment rate) of COVID-19 cases in mainland China was about 6.9% (95%CI: 3.5 – 14.6%) before 29^th^ January 2020, and then increased to 41.5% (95%CI: 30.6 – 65.1%) afterwards. The early reporting rate is very low and about 93.1% of all infections were undetected prior to 29^th^ January 2020 (c.f.^7,9,10^), which might be mainly due to the limited knowledge and unclear definition of the novel disease^11^. The reporting rate of 41.5% at the late stage reflects the increased awareness of the virus and consequently escalating rate of medical help seeking behaviour for respiratory symptoms; however, this rate appears not high and might indicate that many infections are of mild symptoms or asymptomatic. China CDC^12^ used data up to 11^th^ Feb 2020 and estimated that COVID-19 has been mild for 81%. The high proportion of mild symptomatic or asymptomatic infections was further confirmed by a well-investigated outbreak on Diamond Princess cruise ship during February 2020: among 696 confirmed cases, 410 (58.9%; c.f.^13^) are asymptomatic^14^.

**Table 2.**
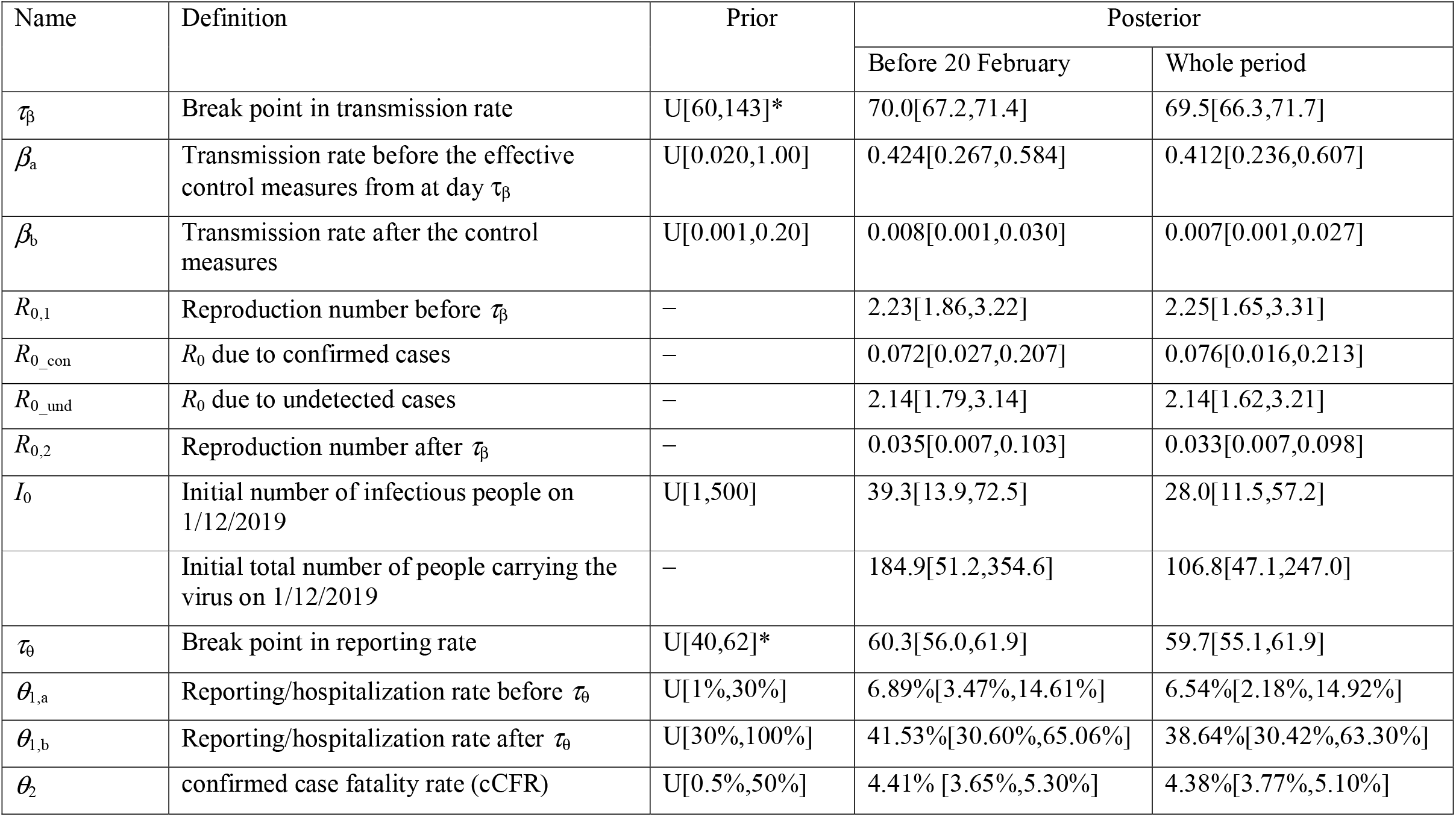
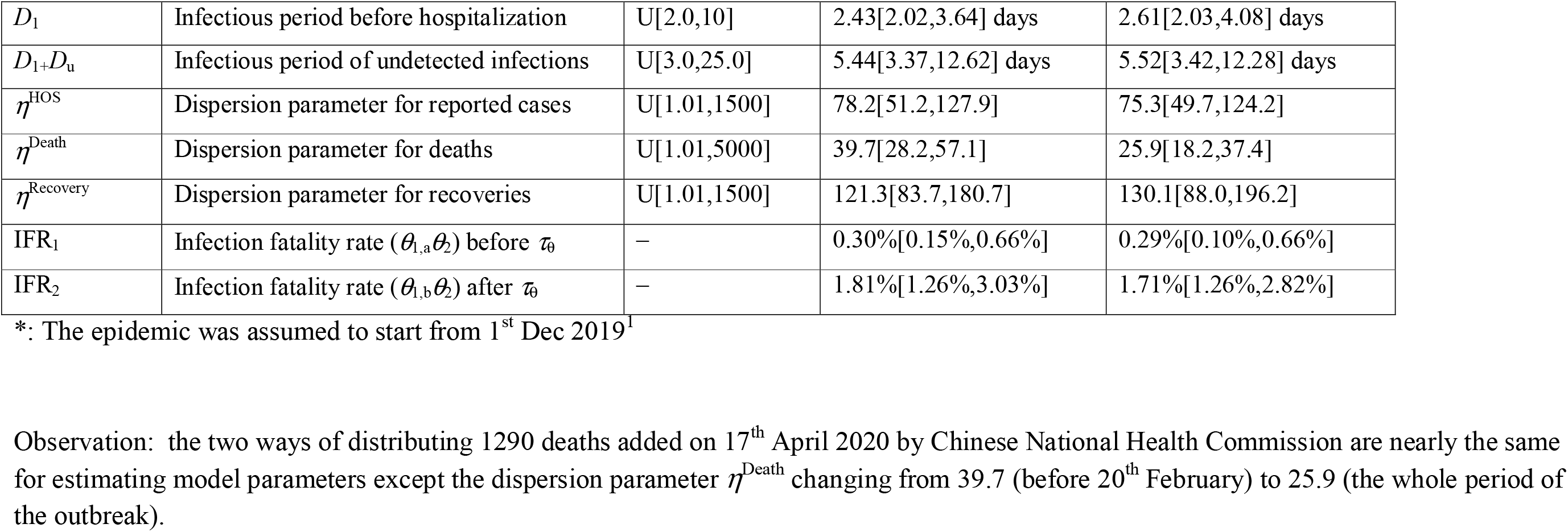
Estimates of model parameters for the COVID-19 outbreak within mainland China. The 1290 deaths within Hubei province added on 17^th^ April 2020 were distributed over either the period before 17^th^ April (i.e., the whole period) or before 20^th^ February 2020 in proportional to the daily number of deaths reported before 17^th^ April 2020. The relative infectivity of undetected cases *ξ* = 1 (i.e., both undetected and confirmed infections are of the same infectivity).

At the early stage before 8^th^ February 2020, the transmissibility of COVID-19 is high with *R*_0_ = 2.23 (95% confidence interval (CI): 1.86 – 3.22); However, it reduced dramatically to 0.04 (95%CI: 0.01 – 0.10) from 8^th^ February 2020 (95% CI: 5^th^ – 9^th^ February 2020). This estimate of *R*_0_ at the early stage are consistent with most of previous estimates ^3,6,7,8,9,10,15–20^. The integrated intervention measures is estimated to reduce transmissibility ot 1.8% of its initial value (c.f.^3^). In our model, the basic reproduction number is contributed by two parts: the confirmed cases and undetected infections. By fixing the duration from onset of symptoms to death and recovery at 17.8 days and 22.6 days respectively^21^, the infectious period (*D*_1_) for these that are symptomatic and confirmed and then quarantined is 2.43 days (95%CI: 2.02 – 3.64 days) and the infectious period for undetected infections (*D*_1_+*D*_u_) is 5.44 days (95%CI: 3.37 –12.62 days). Our estimate of infectious period before isolation is comparable with the estimates of ^22-25^; though it appears to be shorter than that obtained by^3^: 5.19 days (95%CI: 4.51 – 5.86 days). The estimate of infectious period for the infections that were not detected and not quarantined is in agreement with the reviewed range^26,27^ from 1 to 14 days.

Both confirmed cases and undetected infections are assumed to be of the same infectivity in this study; combining with the estimate that only about 6.9% of infections were confirmed at the early stage, this suggests that at the early stage, about 96% of *R*_0_ (Table 2) is due to the undetected infections. If the infectivity of undetected infections is only half or one third of that of confirmed cases^7^, the ascertainment rate increases to 12.2% or 17.6% (Supplementary Table S2.1). And under the conditions where undetected infection are less infectious, the 88% or 71% of *R*_0_ is due to the undetected infections. This indicates that only isolating confirmed cases and their contacts is not enough to stop the spread and that the main factor that stopped the COVID-19 outbreak in mainland China was the dramatic drop in contact rates among the general population (c.f.^7^). This may further explain why the impact of the draconian control measures executed in China since the 23^rd^ January 2020 took about 16 days (i.e., delay from 23^rd^ January to 8th February 2020) to effect in stopping the transmission within community.

Our model analysis shows the confirmed case fatality rate (cCFR) is 4.41% (95%CI: 3.65 – 5.30%). This is slightly below the estimate of ^28^: 5.65% (95%CI: 5.50–5.81%). Deng et al^28^ used individual information of cases to obtain their estimate which is close to both the corresponding crude or naïve confirmed case fatality risk: 4632/82758 = 5.60% and the approximator of deaths/(deaths + recoveries) = 4632/ (4632+78112) = 5.60% as of 21^st^ April 2020. Based on the data up to 11^th^ February 2020, Verity et al^21^ suggested that the overall CFR for China outbreak is 1.38% (95%CI: 1.23-1.53%). Wu et al^29^ found the overall symptomatic case fatality rate (the probability of dying after developing symptom) is 1.4% (95%CI: 0.9–2.1%). Russell et al^30^ based on naive estimates to obtain CFR in China to be 1.2% (95% CI: 0.3–2.7). Our estimate of CFR is higher than these. This should be obvious because these three studies did not include the 1290 deaths added on 17^th^ April 2020 by Chinese National Health Commission^31^. Within the Synthesis model, the hidden transmission dynamics was inferred through observables associated with the disease and so was the number of total infections during the outbreak in mainland China: we found that there were 238894 (95%CI: 127486 – 387917) infected people in mainland China up to 21^st^ April 2020 (Figure 2). Using this information, our estimate of infection fatality rate (IFR) is 0.30% (95%CI: 0.15 – 0.66%) prior to 29^th^ January 2020, and it increased to 1.81% (95%CI: 1.26 – 3.03%) afterwards. This is comparable with estimates of ^21,30^ since both studies were performed before the addition of 1290 deaths on 17^th^ April in Wuhan.

**Figure 2.**
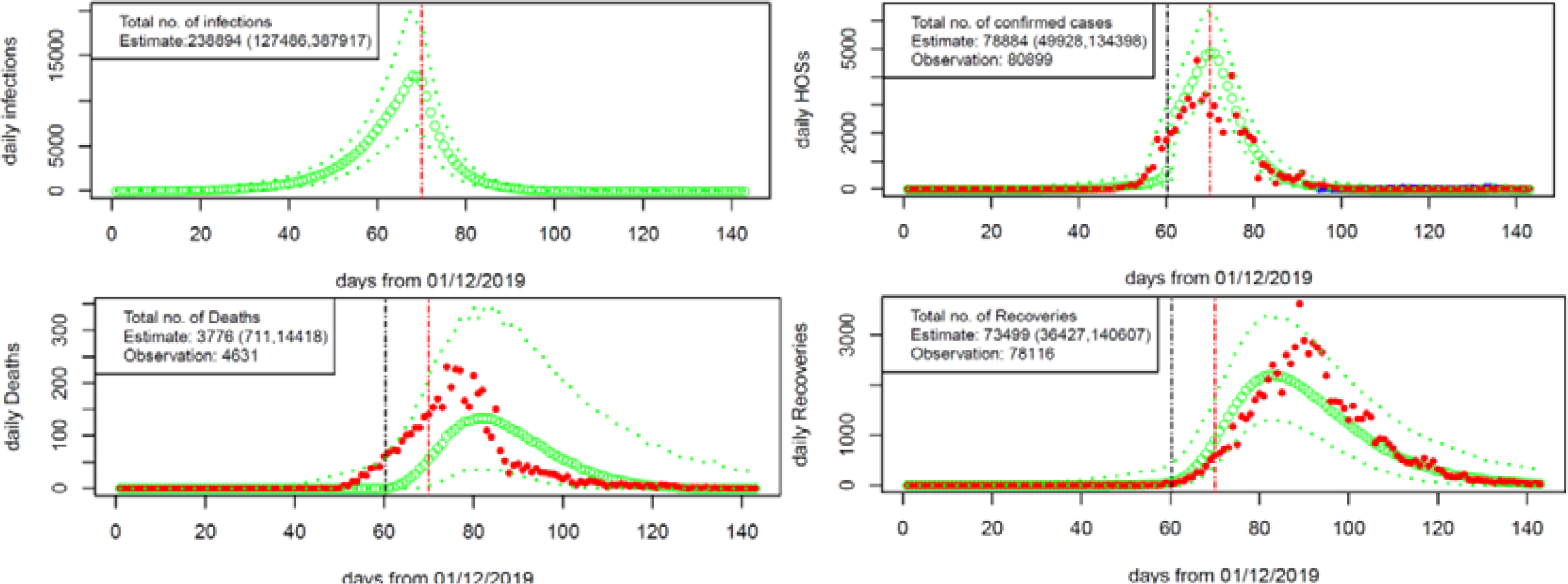
Epidemic curves from 1^st^ December 2019 to 21^st^ April 2020. Model prediction of infections (top left panel) and model fitting to the observations of confirmed cases (top right panel), deaths (bottom left panel) and recoveries (bottom right panel) are shown. The model predictions are obtained under the redistribution of 1290 death added on 17^th^ April 2020 to the period before 20^th^ February 2020 in accordance to the daily number of deaths reported before 17^th^ April 2020. The green dotted lines represent the model predictions (large green circles for median and thin green dotted lines for lower and upper levels of 95%confidence interval). The red points are the observed data. The red vertical line denotes the estimate of break point in transmission rate and the black vertical line the break point in detection and reporting rate. In top right panel, the blue points represent daily number of imported cases. Note that the daily number of confirmed cases (15152) on 12^th^ February 2020 (day 74) is beyond the range shown in the top right panel.

Figure 2 shows the model fitting to the daily number of hospitalizations (confirmed cases), the daily number of deaths, and daily number of recoveries. Our theoretical model well reconstructs the dynamic changes in daily numbers of confirmed cases and recoveries. However, the model predictions of the daily number of deaths shifted about one week late. Our model inference suggests that on 1^st^ December 2019, there were 185 (95%CI: 51 – 354) people who carried SARS-CoV-2 virus. If the public were made aware of COVID-19 earlier and the control measures started earlier, the size of outbreak should be kept much smaller. This can be quantitatively analysed by assuming the same epidemiological characteristics but moving the start time of both reporting and transmission turning time points. The results are listed in Table 3. It shows that if the control measures started one week, two weeks, or three weeks earlier, about 51% (60%), 76% (86%)and 89%(96%) of the confirmed cases (and deaths) would be averted. However, if the control measures started one week, two weeks, or three weeks late, the numbers of the confirmed cases (and deaths) would increase 2.0-fold (2.2), 4.1-fold (4.7) and 8.3-fold (9.76) across mainland China, respectively. This estimate of the effect of start time of control measures is smaller than but comparable with what Lai et al^2^ and Yang et al^5^ found, both studies estimated only the number of infections.

**Table 3.** Impact of start time of control measures during the outbreak in China. Here the total numbers of different cases from 1^st^ December 2019 to 21^st^ April 2020 were shown under different start dates of control measures.

| Start date | Number of infections | Number of confirmed cases | Number of deaths |
| --- | --- | --- | --- |
| 2<sup>nd</sup> Jan 2020 | 27036[8296, 61744] | 8556[2433, 25656] | 129[0, 3914] |
| 9<sup>th</sup> Jan 2020 | 56037[22404, 108590] | 18296[7566, 42499] | 527[0, 6042] |
| 16<sup>th</sup> Jan 2020 | 115065[55009, 197000] | 38126[20377, 72772] | 1512[90, 9279] |
| 23<sup>rd</sup> Jan 2020 | 238894[127486, 387917] | 78884[49928, 134398] | 3776[711, 14418] |
| 30<sup>th</sup> Jan 2020* | 474519[271147, 730166] | 161119[111010,241228] | 8292[2578, 22777] |
| 6<sup>th</sup> Feb 2020* | 957486[564094, 1489883] | 326008[231093,482038] | 17689[7580, 37699] |
| 13<sup>th</sup> Feb 2020* | 1926104[1145812,3128808] | 656380[458844,1009816] | 36314[18731,66699] |

\* Noted: As from 25<sup>th</sup> January 2020, China will start its Chinese New Year holidays, the contacts between people become much higher than the other periods of the year, and hence the estimated numbers of cases here are only conservative.

To compare how the epidemic within the epicentre differs from that over the whole nation, we also obtain the estimates of model parameters for Hubei province (Supplementary Table S3.1). The results show that within the epicentre, transmissibility is only slightly higher (*R*_0_ = 2.34 (95%CI: 1.92 – 3.55) versus 2.23 (95%CI: 1.86 – 3.22)). Ascertainment rate at the early stage within Hubei province is more than double the national rate (15.4% (95%CI: 6.8 – 28.7%) versus 6.9% (95%CI: 3.5–14.6%)); whereas the amount of transmission due to undetected cases is slightly reduced (93% versus 96%). The confirmed case fatality rate is higher (cCFR= 5.16% (95%CI: 4.21–6.28%) in Hubei province versus 4.41% (95%CI: 3.65– 5.30%) over the whole nation with the infection fatality rate at early stage of the outbreak being more than double the national rate (0.79% (95%CI: 0.35–1.53%) versus 0.30% (95%CI: 0.15–0.66%).

## Discussion

Understanding the transmissibility and severity of the novel coronavirus (SARS-CoV-2) is paramount; further disclosing how its rapid spread was brought under control in mainland China is of practical implication for other countries now facing the ongoing outbreaks of COVID-19. In this study we have explored these by using a mathematical model to reconstruct the COVID-19 outbreak in mainland China from 1^st^ December 2019 to 21^st^ April 2020 and assess the impact of its unprecedented control measures. Our analyses indicate that the SARS-CoV-2 has a basic reproduction number of 2.23 under the situation of no intervention measures and therefore is highly transmissible. The fatality rate among those that are symptomatic and confirmed is about 4.41%. The draconian control measures taken by Chinese government from 23^rd^ January 2020 has brought the spread of SARS-CoV-2 under control in mainland China. However, it took more than two weeks for the effect of control measures to emerge.

The massive vigorous actions taken by the Chinese government have stopped the spread of COVID-19 in mainland China. This was achieved under very strict control measures, which dropped the transmissibility from *R*_0_= 2.23 to 0.04. We found that the infectious period before isolation for the confirmed cases is about 2.4 days and the infectious period for these undetected is about 5.4 days ranging from 3.4 to 12.6 days. Although increasing the rate of detection and quarantine of symptomatic cases can help reduce the sources of infection (c.f.^2^), the main force of transmission is from the undetected cases which contributed the vast majority of the transmissibility of SARS-CoV-2 during the early stage of the outbreak in mainland China (c.f.^7^). Hence it is the restriction measures, which limited the mobility of the general population that stopped the spread of SARS-CoV-2 virus in mainland China. Through model fitting to observed data in mainland China, we found that the model predicted fourteen times higher cases than were reported at the early stage of the outbreak; even at the late stage since 29^th^ January 2020, the estimated number of infected people were more than twice that were reported. Based on the epidemiological characteristics obtained from COVID-19 outbreak data, our analyses suggest that even if the control measures started one week earlier, this will avoid 51% confirmed cases and 60% deaths. If starting three weeks earlier, then 89% confirmed cases and 96% of deaths will be averted (c.f.^2^). This reinforces our common sense: the earlier discovering and earlier response, the easier and better control strategy.

As in ^6^, we assumed the latent period is equal to the incubation period. In view of evidence that there is pre-symptomatic transmission^27,32,33^, it is interesting to know whether this will alter the results of our model. For this, we modified the model system (Figure 1) by dividing exposure stage *E* into two equal sub-classes *E*_1_ and *E*_2_ and assuming people in *E*_2_ can transmit the virus with the same infectivity of ill cases (SI Section 4). We obtain the quite similar results (Supplementary Table S4.1): For example, the basic reproduction number before the control measures is 2.24 (95%CI: 1.86 – 2.88) among which 95% were due to undetected cases, and the cCFR is 4.41% (95%CI: 4.41 (95%CI: 3.67 – 5.37%). This indicates our model is robust to possible pre-symptomatic transmission. Nevertheless, this might imply that population models of SARS-CoV-2 virus transmission cannot distinguish where or not there is pre-symptomatic transmission.

The non-pharmaceutical interventions executed in mainland China have stopped the spread of the virus, but the risk of another potential outbreak lies in the nearby future. This argument come straightforward^3^. As our model estimated, at most 238894 individuals got infected up to 21^st^ April 2020 during the outbreak, which is only 0.02% of the Chinese population. Even if all those recovered from infection have built complete immunity^34^, this immunity level in Chinese population is too lower than the herd immunity of 1-1/*R*_0_ > 50% which is required to protect the population^35,36^. This simply implies that once the strict quarantine measures currently executed in mainland China is released, the rebound of COVID-19 in China is very likely especially under the current rising outbreaks across the rest of the world (c.f.^37^). Nevertheless, the non-pharmaceutical interventions have halted the spread of SARS-CoV-2 virus in mainland China and bought time for vaccines and drugs to be developed and used late on.

It is of interest to compare the novel coronavirus SARS-CoV-2 with other two coronaviruses: SARS-CoV-1 and MERS-CoV that caused large outbreaks in human populations. SARS-CoV-1 has *R*_0_ from 2–5^38^ and case-fatality rate of 9.6% among probable cases in mainland China^39^. MERS-CoV in the 2015 outbreak in South Korea has been estimated to have a *R*_0_ from 2–7^40^ and case fatality rate of 34.5% among laboratory-confirmed cases^41^. It indicates that SARS-CoV-2 is nearly as transmissible as SARS-CoV-1 and MERS-CoV, but less severe. Furthermore, there was no evidence of a super-spreader event occurring in any of the Chinese health facilities serving COVID-19 patients, which is distinct from the 2003 outbreak of SARS-CoV and 2015 MERS-CoV outbreak in South Korea. Unfortunately, due to the advanced modern transportation, people can move easily and quickly across the world, and the SARS-CoV-2 quickly spread to other countries. This is in a sharp contrast with both SARS-CoV-1 and MERS-CoV which have been controlled and confined within relatively limited areas of the world. This sharp difference may be attributed to another aspect of the coronaviruses: a large proportion of SARS-CoV-2 infections are of mild symptoms or asymptomatic^12-14^, while both SARS-CoV-1 and MERS_CoV are highly symptomatic^20,32^. This characteristic of SARS-CoV-2 virus will make it a real challenge for humans to control and manage it.

The strength of our analysis comparing with previous studies^2,3,5,6,20^ lies in two aspects: our investigations are based on the three datasets (confirmed cases, death and recovery) and we model the outbreak over a long period (143 days) which should avoid any bias and confounding arising due to observations over a short period. This study also has several limitations. To model complicated processes of transmission dynamics and disease reporting, Synthesis model has been simplified in several aspects. To reflect the temporal change in both ascertainment rate and transmissibility of COVID-19, two different values are assumed for each of them. The change in ascertainment rate and transmissibility may gradually take place during the outbreak as do the public awareness and interventions^3^. For example, Tsang et al^11^ found the ascertainment rate changed as the case definition for COVID-19 changed from initially narrow to gradually wider during the period from 15^th^ January to 3^rd^ March. In the current study, we assume the confirmed case fatality rate (cCFR) remained unchanged during the outbreak in mainland China. It may reduce with time as medical conditions and clinical treatments improved. The time-to-event intervals such as the delay from symptom onset to death may also changes as epidemic grows^21,23,25^. Further, in this study we ignore the heterogeneity in both geography and age ^3,7,12^. To provide more specific and practically useful information for control measures, it needs to look at variation in regions^3,7^ and age groups^12^. A further limit is we model the overall effectiveness of integrated intervention measures rather than the different types of control measures and therefore cannot provide specific information for their relative impacts on stopping the spread of infection (c.f.^2,3,15^).

In conclusion, our finding that the main driver of transmission of SARS-CoV-2 at the early stage of the outbreak in mainland China came from the undetected infections provides vital information for policy makers when designing the optimal intervention strategies. Under the situation where vaccine and effective drugs are not available, early detection and isolation are essential for containing and controlling the spread of SARS-CoV-2 but the most crucial is to reduce contact rates among people in the general population.

## Methods

### Data

We extracted the following data relating to COVID-19 for mainland China from 3 datasets on the website of Chinese National Health Commission: the daily number of confirmed cases who were confirmed / admitted to hospital, the daily number of deaths and the number of patients who had recovered each day. The originally reported data from 1^st^ December 2019 to 21^st^ April 2020 are collected for this study. Here the data are given by the symptom onset date during the period 1^st^ December 2019 – 1^st^ January 2020 from^1^ and by reporting date thereafter from the website of Chinese National Health Commission. The reporting date is assumed to be the same as the date that cases were diagnosed. Before 12^th^ February 2020, confirmed cases were defined as those who were positive for SARS-CoV-2 on PCR; from 12^th^ February 2020 confirmed cases were defined (for the epicentre Hubei province) as those who were either clinically diagnosed or positive for SARS-CoV-2 on PCR^11^.

On 17^th^ April 2020, Chinese National Health Commission raised the total death number in Wuhan city from 2579 to 3869^31^. The announcement of the Chinese National Health Commission stated that these added 1290 deaths most likely occurred before 20^th^ February 2020. Because, after this date, the number of hospitals that can treat COVID-19 patients increased from 2 to 48, and the HOUSHEN and LEISHEN hospitals, and FANGCANG shelters that have been constructed since also provided much more beds meeting the needs of COVID-19 patients with different symptoms. Further, the system of data collection improved rapidly, and the number of missed cases/deaths decreased greatly. As the detailed information for these 1290 deaths is not available, for the sake of model fitting these 1290 deaths were relocated in the proportion to the number of deaths on each day reported before from the date of first death (10^th^ January 2020) to 20^th^ February 2020. For example, the number of deaths reported on 18^th^ January 2020 was 26, the number is now corrected to 26+26×1290/2236 = 26+15 =41. Here 2236 is the cumulative number of death up to 20^th^ February 2020. The so-modified death data will be used in this study. For sensitivity analysis, we also assume that these 1290 deaths added on 17^th^ April 2020 were distributed in the proportion to the daily number of deaths reported before over the entire period from 10^th^ January to 17^th^ April 2020. The results shown in Table 2 are very similar under two ways of relocating 1290 deaths added on 17^th^ April 2020.

### Model

In this study we use a synthesis model^42^ (Figure 1), which combines the hidden transmission dynamics of SARS-CoV-2 virus and the reporting system of COVID-19, to investigate the transmissibility and severity of COVID-19 and the efforts to contain and control the spread of SARS-CoV-2. We assume the transmission dynamics of SARS-CoV-2 virus is described by an SEIR compartmental model. That is, a susceptible person (*S*) can contract SARS-CoV-2 virus from infectious persons and then enter the latent class (*E*); after a latent period (*L*), the exposed person progresses to become infectious (*I*) for a period and can transmit the SAR-CoV-2 virus before recovering or dying (*R*). We assume that some of infected people (with severe symptoms) will be detected and admit to hospital and will then be treated and isolated from the community; while other mild or asymptomatic infections (*I*_u_) will not be detected and hence can continue to exist in the community as sources of infection. For simplicity, we assume that the latent period is equal to the incubation period, which is fixed at the value of *L* = 5.2 days as estimated by^17,25,43^. SARS-CoV-2 virus can be transmitted by three possible modes: respiratory transmission (through respiratory droplets when symptomatic people sneeze or cough), aerosol transmission (through fine virus particles that were aerosolized) and contact transmission (through contacting the contaminated surface). In view of this easy and quick mode of transmission, random mix among people is a reasonable assumption. Although China is a large country with a population of 1400050000 people residing on a huge area of 9,596,960 km^2^, the recent urbanization and the development of rapid transport systems in China make it easy and quick for its people to move around the country. Hence it should be rational to model the transmission of COVID-19 within the whole country as a well mixing population. For comparison, we also model the spread of COVID-19 within Hubei province where the outbreak started (SI Section 3).

The synthesis model is approximated by equation (1).

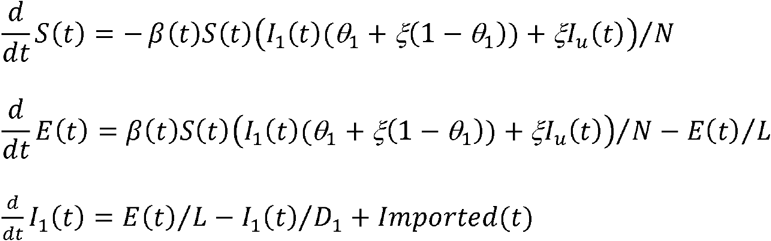

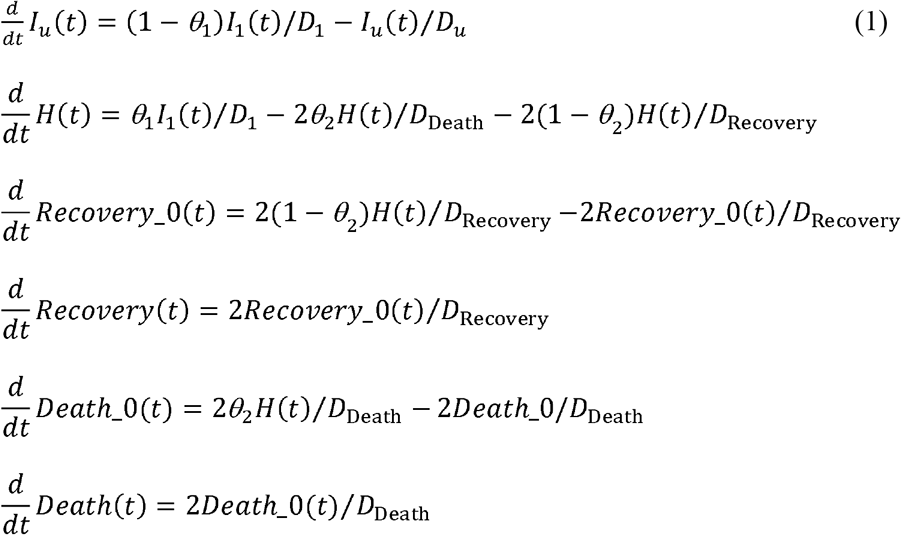

Here *N* =1400050000 is the size of total population in mainland China and is assumed to be constant during the outbreak. The 12 compartments are defined in Table 4 and the definitions of model parameters are given in Table 2. Note that an item *Imported*(*t*) in equation for *I*_1_ is included to account for the imported cases which come back from other countries to China since 24^th^ March 2020.

**Table 4.** Definitions of model compartments

| Name | Definition |
| --- | --- |
| **Transmission dynamics** |  |
| $S(t)$ | Number of susceptible people at time $t$ |
| $E(t)$ | Number of exposed (infected but not yet infectious) people at time $t$ |
| $I_1(t)$ | Number of all infectious people at time $t$ who have not yet been detected |
| $I_u(t)$ | Number of infectious people at time $t$ who have not been detected and will remain undetected |
| **Disease reporting** |  |
| $H(t)$ | Number of people who were hospitalized/reported due to COVID-19 at time $t$ |
| $Death\_0(t)$ | Number of people in the “Death_0” compartment (between $H$ and $Death$) at time $t$. |
| $Death(t)$ | Number of people who have died from COVID-19 at time $t$ |
| $Recovery\_0(t)$ | Number of people in the “Recovery_0” compartment (between $H$ and $Recovery$) at time $t$ |
| $Recovery(t)$ | Number of recovered people at time $t$ |

As a novel virus, no effective pharmaceutical interventions were available. Non-pharmaceutical interventions are therefore essential public health response to the outbreaks. These include isolating ill persons, contact tracing, quarantine of exposed persons, travel restrictions, school and workplace closures, and cancellation of mass gathering events. These measures were expected to reduce transmission^2,5^. During the period of COVID-19 outbreak in China, the day, 20^th^ January 2020, when the information that “SARS-CoV-2 virus can transmit among people” was announced in public marked some huge and quick changes in people contact behaviours and rates. These changes were further strengthened when the lockdown of epicentre Wuhan city from 23^rd^ January 2020 and then quick follow-up of nationwide restrictions of mobility were enforced (Table 1 Timeline of the outbreak in China). To model the potential changes in transmission rate due to these combined and dramatic control measures taken by Chinese government, we simply assume there existed a turning point in time *τ*_β_ such that the transmission coefficient varies as shown in the equation (2)

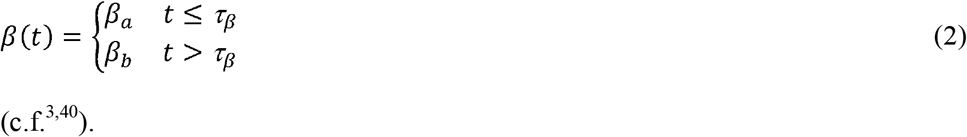

At the early stage of COVID-19 outbreak in China, due to the limited knowledge and unclear definition of COVID-19, the rate of detection and reporting was very low (i.e., under-detection and under-reporting). However, the rate of detection and reporting increased quickly with the knowledge about the COVID-19 and availability of advanced techniques to test SARS-CoV-2^11^. Especially since 20^th^ January 2020, searches for cases, diagnosis and reporting have sped up. Local governments across China encouraged and supported routine screening and quarantine of travellers from Hubei Province to discover COVID-19 infections as early as possible^2^. To reflect the changes in detection and reporting of COVID-19 cases around 20^th^ January 2020 when the SAR-CoV-2 was publicly declared to able to transmit among people, we assume the proportion of detection (or rate of ascertainment) varied with time as

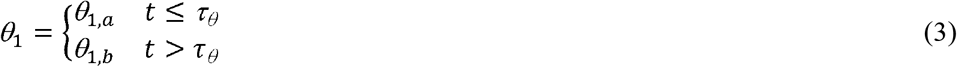

Here τ_θ_ is the turning point in detection and reporting.

In the disease reporting system of the synthesis model (see the shaded part of Figure 2), we assume all confirmed cases were hospitalised. The hospitalised patients either died at the probability of *θ*_2_ or discharged because of recovery. Despite sporadic cases of COVID-19 that might not have chances to be hospitalized and died at home especially at the early stage of the outbreak, our assumed flow of confirmed COVID-19 patients should well approximate actual procedures during the outbreak in mainland China^44^. In this study we fix the durations from onset of symptoms to death and to hospital discharge at (*D*_1_+*D*_eath_ =) 17.8 days and (*D*_1_+*D*_Recovery_ =) 22.6 days, respectively, the estimates of ^21^ from the China outbreak data. To allow the durations from hospitalization to death and from hospitalization to recovery to have gamma distribution rather than the usual exponential distribution, we introduce the in between compartments *Death*_0, and *Recovery*_0. As we use (mostly) data of reported dates, the time-event-length (duration) should be thought to include such reporting delay, which is not explicitly treated in this study. Fortunately, modelling technique can code with such external noise^6^.

The basic reproduction number *R*_0_, which was defined as an average number of secondary infections generated by an infectious person introduced into a completely susceptible population, is an important quantity to characterise the transmissibility of infectious agents^35^. At the early stage of our model system, it is given by

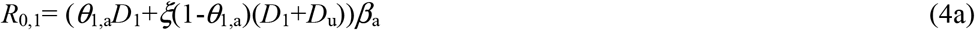

while at the late stage after the time point defined as max(*τ*_β_, *τ*_θ_),

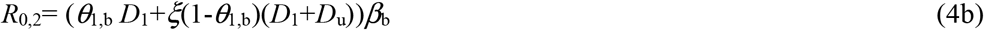

From equation (4a), the contribution from undetected infections to the transmissibility at the early stage is

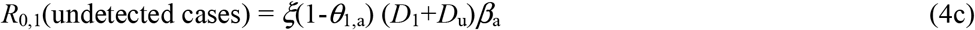

Here *D*_1_+*D*_u_ is the total infectious period of the infected people who were not detected. The parameter *ξ* is introduced to measure the relative infectivity of undetected infections to confirmed cases, and it will take the value of 1.0 (i.e., both undetected and confirmed infection are of the same infectivity). To see how our results may change with different relative infectivity^7^ the results for situations of *ξ* = 1/2, 1/3 will be given in Supplementary Table S2.1.

#### Initial seeding

Wu et al^20^ Lancet assumed the epidemic during 1^st^ – 31^st^ December 2019 was seeded by constant zoonotic force of infection that caused 86 cases (twice the 43 confirmed cases with zoonotic exposure) per day before market closure on 1^st^ January 2020. Kucharski et al^6^ assumed the outbreak started with a single infectious case or 10 cases on 22^nd^ November 2019. In this study we assume the epidemic started from 1^st^ December 2019 with initial infections of *I*_1_(0) = *I*_0_ which is to be estimated from model fitting to data. (The detail of how infections were seeded is given in SI Section1)

### Inference method

We denote the model parameter to be inferred as Θ = {*I*_0_, βa, β_b_, *τ*_β_, *θ*_1,a_, *θ*_1,b_, *τ*_θ_, *θ*_2_, *D*_1_, *D*_u_} which is listed in Table 2. For each set of parameter values, the Runge-Kutta fourth order method was used to solve the model equations and to obtain model predicted time series of infections, confirmed cases, deaths, and recoveries. In the inference of model parameters, directly observed dataset of confirmed/hospitalized/reported cases (denoting it as HOS for short in the following), Death and Recovery are used as illustrated in the following. To capture the large dispersion in the daily numbers of these cases, the negative binomial likelihood function was assumed. The likelihood for observed number *x*^C^ (*t*) of cases on day *t* is given as

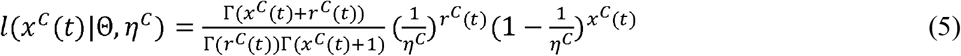

where

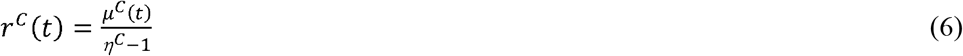

Here *η*^*C*^ is the dispersion parameter and *μ*^C^(*t*) are the predictions of the cases on day *t* from Synthesis model (1). Superscript *C* represents three different datasets: HOS, Death and Recovery.

Special attention was paid to the extra high daily number of cases (15152) on 12^th^ February 2020 (day 74 from 1^st^ December 2019) due to the change in the case definition in Hubei province^11,45^. In principle, these cases might have been accumulated over the days before 12^th^ February 2020. To avoid the complexity, only the cumulative numbers of cases on the 12^th^ February and daily numbers of cases after that will be used in the model inference. Let the reported daily number of HOS be represented by *x*(1), *x*(2), …, *x*(*T*), with *T* being the number of days from 1^st^ December 2019 to 21^st^ April 2020. The likelihood of the cumulative number of cases on 12^th^ February 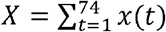 is assumed to be

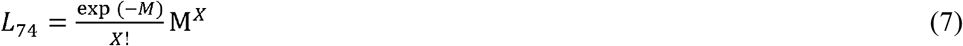

Here 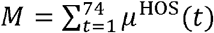 represents the cumulative number of confirmed cases predicted by model. Assuming that the observational daily number of death: *y*(1), *y*(2),…, *y*(*T*) and daily number of recovery: *z*(1), *z*(2),…, *z*(*T*) are conditionally independent, the total likelihood given model parameters Θ is

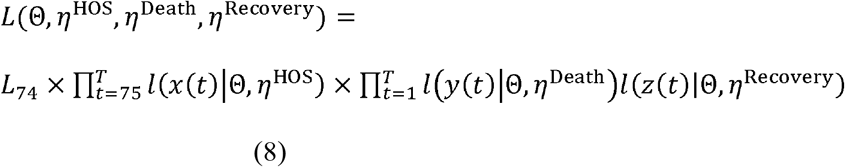

#### Parameter inference

We *as*sume the uninformative prior distributions *f*(Θ) which are uniform for parameters (Table 2). Employing Bayesian framework through the combination of the prior distribution *f*(Θ) and the likelihood *L*(Θ,*η*^HOS^, *η*^Death^, *η*^Recovery^ ***x, y, z***), the posterior distribution can be obtained by Markov Chain Monte Carlo simulations (MCMC)^46^ (Birrell et al 2011). From these samples, we can obtain medians and their 95% confidence intervals for the model parameters. The posteriors of the model parameters will provide the estimates of the transmissibility and the severity of SARS-CoV-2 in mainland China and the effects on transmissibility of control measures executed by Chinese government.

## Data Availability

All the data are given in the paper

## Acknowledgements

This work is jointly supported by Public Health England, UK and Innovative Research Team of Yunnan Province (2019(6)), China.

## Author Contributions

Conceived and designed the mathematical model and analysis: X.-S.Z., E.V., A.C., D.D.A. and W. L.; collated data: Z.C.; performed the study: X.-S.Z. and W.L.; wrote the paper: X.-S.Z. and E.V.

